# Impact of cattle on joint dynamics and disease burden of Japanese encephalitis and leptospirosis

**DOI:** 10.1101/2020.06.22.20137364

**Authors:** Mondal Hasan Zahid, Christopher M. Kribs

## Abstract

Japanese encephalitis (JE) is a mosquito-borne neglected tropical disease. JE is mostly found in rural areas where people usually keep cattle at home for their needs. Cattle in households reduce JE virus infections since they distract vectors and act as a dead-end host for the virus. However, the presence of cattle introduces risk of leptospirosis infections in humans. Leptospirosis is a bacterial disease that spreads through direct or indirect contact of urine of the infected cattle. Thus, cattle have both positive and negative impacts on human disease burden. This study uses a mathematical model to study the joint dynamics of these two diseases in the presence of cattle and to identify the net impact of cattle on the annual disease burden in JE-prevalent areas. Analysis indicates that the presence of cattle helps to reduce the overall disease burden in JE-prevalent areas. However, this reduction is dominated by the vector’s feeding pattern. To the best of our knowledge, this is the first study to examine the joint dynamics of JE and leptospirosis.

## 1 Introduction

Japanese encephalitis viral disease (JE) was first documented in 1871 in Japan [1]. Japanese encephalitis (JE) is the main cause of viral encephalitis in many countries of Asia and the western Pacific. 24 countries in the WHO South-East Asia and Western Pacific regions have Japanese encephalitis virus (JEV) transmission risk, which includes more than 3 billion people [1, 2]. JEV is transmitted to humans through bites from infected mosquitoes of the Culex species (mainly *Culex tritaeniorhynchus*). The virus exists in a transmission cycle between mosquitoes, pigs and/or water birds and is transmitted to humans by infected mosquito bite. JE is predominantly found in rural and periurban settings [3, 4]. Each year there are nearly 68,000 clinical cases of JE globally, with approximately 13,600 to 20,400 deaths. JE primarily affects children. Most adults in endemic countries have natural immunity after childhood infection, but individuals of any age may be affected. Most people infected with JE do not have symptoms or have only mild symptoms. However, a small percentage of infected people develop inflammation of the brain (encephalitis), with symptoms including sudden onset of headache, high fever, disorientation, coma, tremors, and convulsions. Approximately 1 in 250 infections results in severe clinical illness. Severe disease is characterized by rapid onset of high fever, headache, neck stiffness, disorientation, coma, seizures, spastic paralysis, and ultimately death. [1, 2]. The case fatality rate for the disease can be as high as 30% among those with disease symptoms. In addition, 20-30% of those who survive suffer permanent neuropsychiatric sequelae [3].

Leptospirosis is a bacterial disease that affects humans and animals. Leptospirosis is considered to be the most widespread zoonotic disease in the world [5]. It is caused by bacteria of the genus *Leptospira* [6]. The bacteria are spread through the urine of infected animals, which can get into water or soil and can survive there for weeks to months. Humans can become infected through contact with urine (or other body fluids, except saliva) from infected animals; also, through contact with water, soil, or food contaminated with the urine of infected animals [7]. In humans, leptospirosis may occur in two phases, where about 10% of infected people move to the 2nd phase [5]. The first phase causes a wide range of symptoms, some of which may be mistaken for other diseases: fever, chills, headache, muscle aches, vomiting, or diarrhea. The second phase, if it occurs, is more severe; the person may have kidney damage, liver failure, meningitis, respiratory distress, and even death [6, 8]. It is estimated that more than 1 million cases occur worldwide annually, including almost 60,000 deaths [8].

In the last couple of decades, many ecological and field studies, along with a few mathematical ones, have been conducted to understand the dynamics of JEV transmission and to find some control measures to reduce JEV prevalence in humans. In 2001, a study used a probabilistic model of pathogen transmission to investigate various control measures for JEV transmission in humans [9]. Outcomes of the study show that a combination of control measures of similar effect (strategies to reduce vector population, **or** strategies to reduce vector-human interactions) is more effective compared to the combination of control measures of different effects (strategy to reduce vector population and strategy to reduce human-vector interaction). In 2014, Khan et al. studied the dynamics of JEV transmission in a pig population in northwest Bangladesh [10]. This study developed an SEIR model to understand transmission dynamics in pigs, and to estimate the potential impact of pig vaccination. Their results found that the prevalence of JE in pig populations can be reduced by up to 89% when 75% of susceptible pigs are vaccinated each year. The next year, in 2015, Lord et al. performed a study to rethink JEV transmission among hosts and vectors [11]. They suggested using a mathematical model parameterized with data to quantify the relative roles of potential species in JEV transmission.

Very little research has been done, to the best of our knowledge, to understand the dynamics of leptospirosis in cattle and humans. In 2017, Chadsuthi et al. investigated the leptospirosis prevalence in livestock and humans in Thailand for 2010-2015 [12]. They tested humans, buffaloes, cattle, pigs, and analyzed collected data. Their analysis found livestock more susceptible to leptospirosis infection compared to humans. Later, in 2018, another study was done to understand the spread of leptospirosis in lambs in New Zealand. Here, researchers used a simple SI model to predict conditions under which the disease would persist in the lamb population [13]. Analysis of this study suggested that increasing the *leptospira* death rate in farms can reduce infection in livestock, and eventually in humans.

There are many countries in Asia where both JE and leptospirosis are prevalent. Taking this reality into account, this study is developed to investigate the prevalence of both diseases in humans. Cattle contribute to leptospirosis infections in humans, acting as a source of *leptospira*. However, the presence of cattle in a domestic or peridomestic setting can be considered as an additional host for JE vectors, where humans act as the primary host. Hence, the presence of cattle increases host richness in the setting, which might eventually reduce the combined burden of these two diseases for humans. Earlier studies showed that an additional host (host richness) in a setting helps to reduce human infections of vector-borne diseases if certain conditions are maintained [16, 17, 18, 20]. Johnson and Thieltges in 2010 identified that host diversity may help to reduce human infections depending on the relative abundance of additional host(s) relative to the focal host [16]. In 2013, Miller and Huppert proved that species diversity in host populations can amplify or can dilute disease prevalence depending on vectors’ preference of host [17]. Recently in 2020, Zahid and Kribs showed that an additional host (dog, reservoir host) in a community can help in reducing the prevalence of visceral leishmaniasis in humans depending on the additional host’s irritability to vector bites [18]. In another study, the same authors established that the presence of an additional host (chicken, incompetent host) in a domestic setting might help to reduce the prevalence of Chagas disease in humans if the distance between the two host populations (humans and chickens) is maintained at a certain range [20].

The presence of cattle helps to reduce human-vector interactions by attracting mosquitoes towards them from humans. Cattle have no contribution to the transmission of JE [21] whereas they act as a source of leptospirosis infections in humans. Thus, in terms of infections, the presence of cattle has two opposite influences on human health – it helps to reduce JE prevalence in humans and contributes to leptospirosis risk in humans. The goal of this study is to understand the dynamics of both diseases in each population, and eventually to understand if the presence of cattle in a setting, where JE and leptospirosis both are prevalent, is helpful to reduce the combined burden of JE and leptospirosis in humans. To answer this question we compare two different peridomestic settings: a setting involving cattle with humans, pigs, and mosquitoes, and the other setting involving humans, pigs, and mosquitoes without cattle. To understand disease dynamics, and to compare the proposed two settings, we develop SIR models for cattle, pig and human populations, and an SI model for the mosquito population. This study estimates the total number of *DALY* s (Disability-Adjusted Life Year) generated from JEV and (in the presence of cattle) leptospirosis infections for both settings and compare them to identify the better scenario. To the best of our knowledge, this is the first work ever to consider JE and leptospirosis infections together. More broadly, this extends the well-studied question in disease ecology of the impact of an additional host to a multi-pathogen context.

## 2 Model development

The goal of this work is to understand the dynamics of Japanese encephalitis virus (JEV) and leptospirosis infections together and to estimate the impact of cattle presence on human infections of these diseases. Therefore, we select a location for this study where both diseases are prevalent. We choose Malkangiri, the southern district of Odisha (Orissa) State in India. Malkangiri has a history of JE outbreaks since 2009 where the cases are documented as Acute Encephalitis Syndrome (AES) [22]. This AES can include infections other than JE. Studies showed that AES cases are not always cases of JE infections, they can be cases of the severe form (the 2nd phase) of leptospirosis infections [23, 24, 25]. This fact, along with the high prevalence of leptospirosis in cattle in neighbor districts [26, 27], illustrates the presence of human leptospirosis cases in our study location.

We use an SIR model to understand the dynamics of JEV among pig, mosquito, and human populations; also, to understand the dynamics of leptospirosis between cattle and human populations. JEV mainly exists in a transmission cycle between mosquitoes and pigs, and humans get the infection of JE from bites of infected mosquitoes (*I*_*m*_) [1, 2]. However, infected humans do not infect feeding mosquitoes due to the lack of sufficient viremia. So, humans are dead-end hosts for JEV [1, 2, 3, 36]. Also, cattle have no role in the maintenance of JEV in nature [21]. Thus, humans and cattle have no contribution to JEV transmission. We consider total number of pig bites as 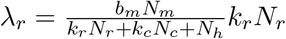 and total number of human bites as 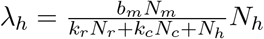,where *N*_*m*_,

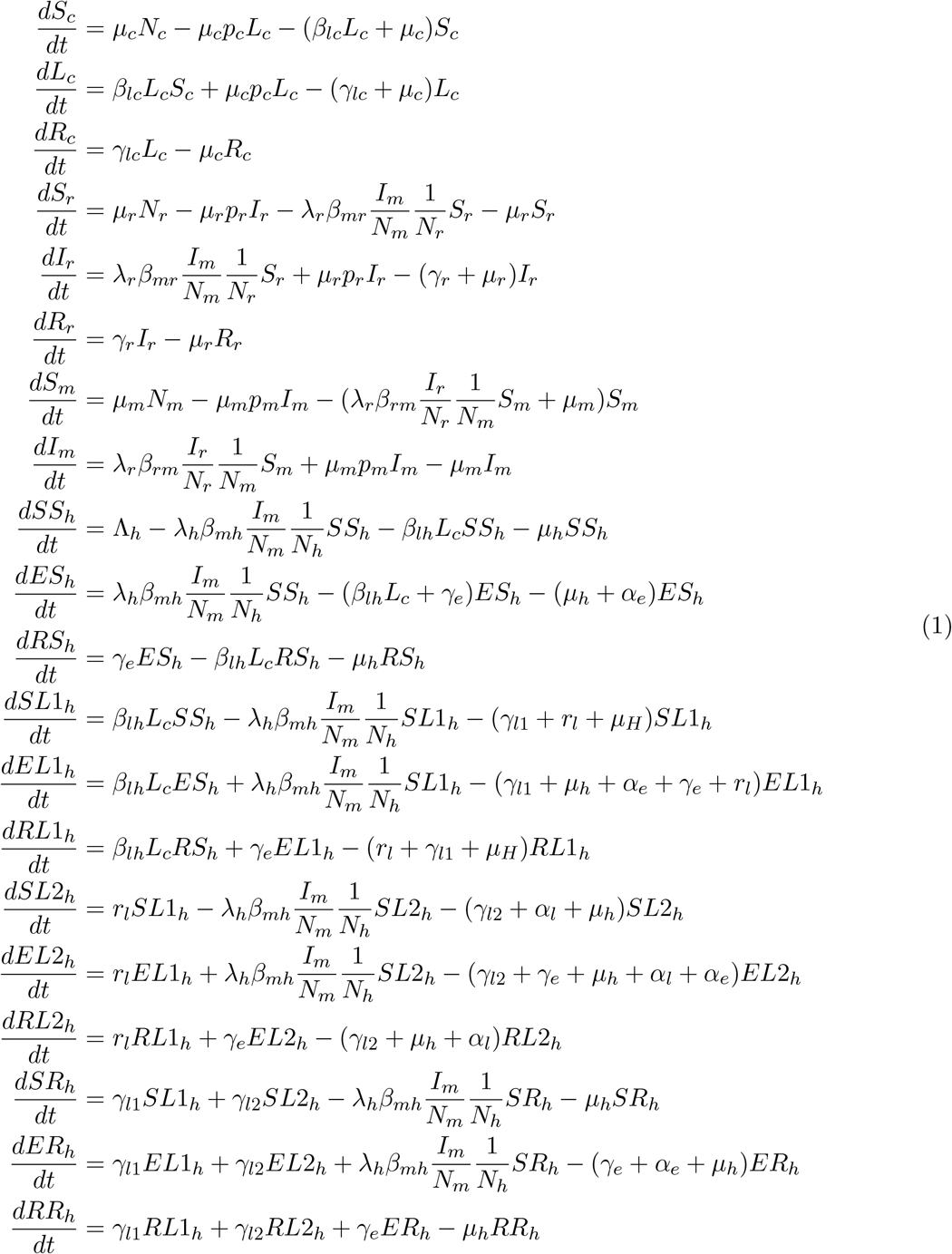

Where

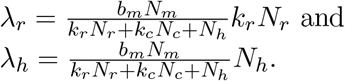

*N*_*r*_, *N*_*c*_, and *N*_*h*_ represent sizes of mosquito, pig, cattle and human populations respectively, *b*_*m*_*N*_*m*_ represents total mosquito bites. Here, *k*_*r*_ and *k*_*c*_ are defined based on mosquitoes’ feeding preference among hosts (see Table 2). In our model development, we consider the vertical transmission of JEV in pigs, and mosquitoes. All these ideas are illustrated in Figures 1, 2, and 3.

**Table 1:**
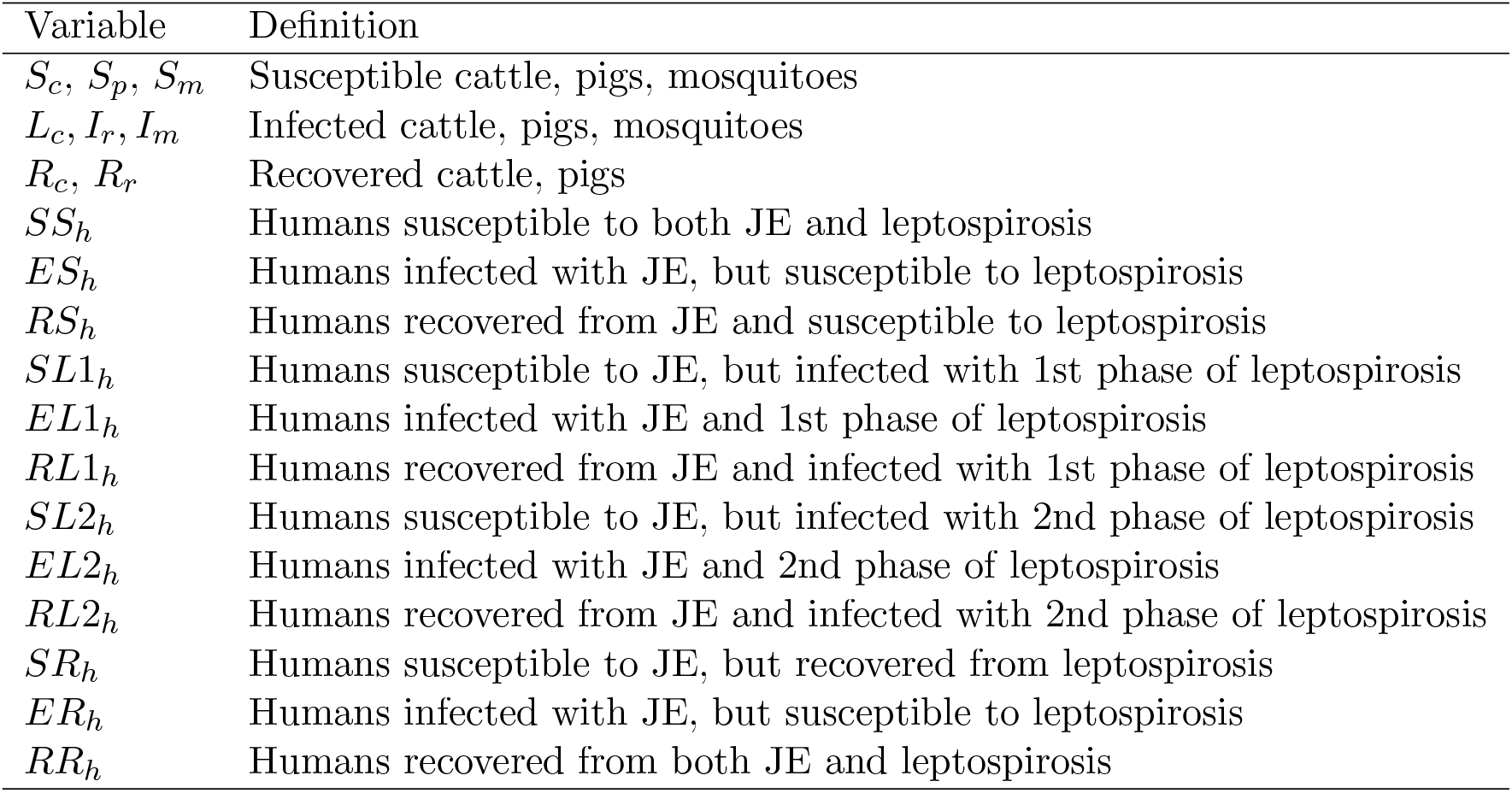
Model variables with definition.

**Table 2:** Summary of model parameters.

| Par. | Definition | Value | Units | Ref. |
| --- | --- | --- | --- | --- |
| $\Lambda_h$ | recruitment rate for humans | 25.87 | human/day | This study |
| $p_c$ | probability of vertical transmissions of leptospirosis in cattle | 0 | - | This study |
| $p_r$ | probability of vertical transmissions of JE in pigs | 0 | - | This study |
| $p_m$ | probability of vertical transmissions of JE in mosquitoes | 0.04 | - | This study |
| $\beta_{lc}$ | infection rate of leptospirosis to cattle | $9.3 \times 10^{-8}$ | 1/day-cattle | This study |
| $\beta_{lh}$ | infection rate of leptospirosis to humans | $9.12 \times 10^{-12}$ | 1/day-cattle | This study |
| $\beta_{rm}$ | infection probability of JE to mosquito from biting pigs | 0.82 | mosquito/bite | [34] |
| $\beta_{mr}$ | infection probability of JE to pigs from mosquitoes' bite | 0.635 | pig/bite | [34] |
| $\beta_{mh}$ | infection probability of JE to humans from mosquitoes' bite | 0.316 | human/bite | [34] |
| $\gamma_e$ | recovery rate from JE | $\frac{1}{7}$ | 1/day | [35] |
| $\gamma_{lc}$ | recovery rate from leptospirosis for cattle | $2.38 \times 10^{-3}$ | 1/day | This study |
| $\gamma_{l1}$ | recovery rate from 1st phase of leptospirosis (humans) | $\frac{1}{10}$ | 1/day | [39] |
| $\gamma_{l2}$ | recovery rate from 2nd phase of leptospirosis (humans) | $\frac{1}{14}$ | 1/day | [39] |
| $r_l$ | transfer rate from phase 1 to phase 2 of leptospirosis | 0.033 | 1/day | This study |
| $\gamma_r$ | recovery rate from JE for pigs | $\frac{1}{4}$ | 1/day | [36] |
| $\mu_c$ | natural death rate for cattle | $3.42 \times 10^{-4}$ | 1/day | This study |
| $\mu_r$ | natural death rate for pigs | $3.91 \times 10^{-4}$ | 1/day | This study |
| $\mu_m$ | natural death rate for mosquitoes | $\frac{1}{59.8}$ | 1/day | [37] |
| $\mu_h$ | natural death rate for humans | $3.96 \times 10^{-5}$ | 1/day | [28] |
| $\alpha_e$ | JE-induced death rate for humans | 0.0383 | 1/day | This study |
| $\alpha_l$ | leptospirosis-induced death rate (during 2nd phase for humans) | $3.13 \times 10^{-2}$ | 1/day | This study |
| $b_m$ | mosquito's biting rate | 0.25 | bite/mosquito-day | [33] |
| $N_c$ | cattle population size of the study area | 302,911 | cattle | This study |
| $N_m$ | mosquito population size of the study area | 68,428,337 | mosquitoes | This study |
| $N_r$ | pig population size of the study area | 37,616 | pigs | This study |
| $N_h$ | human population size of the study area | 653,309 | humans | This study |
| $k_c$ | vector's feeding preference (humans over cattle) | $\frac{46.4}{1.5}$ | human/cattle | This study |
| $k_r$ | vector's feeding preference (humans over pigs) | $\frac{4.8}{1.5}$ | human/pig | This study |
| $\lambda_r$ | encounter rate between pigs and mosquitoes | 142,102 | bite (total)/day | This study |
| $\lambda_h$ | encounter rate between humans and mosquitoes | 771,250 | bite (total)/day | This study |

**Figure 1:**
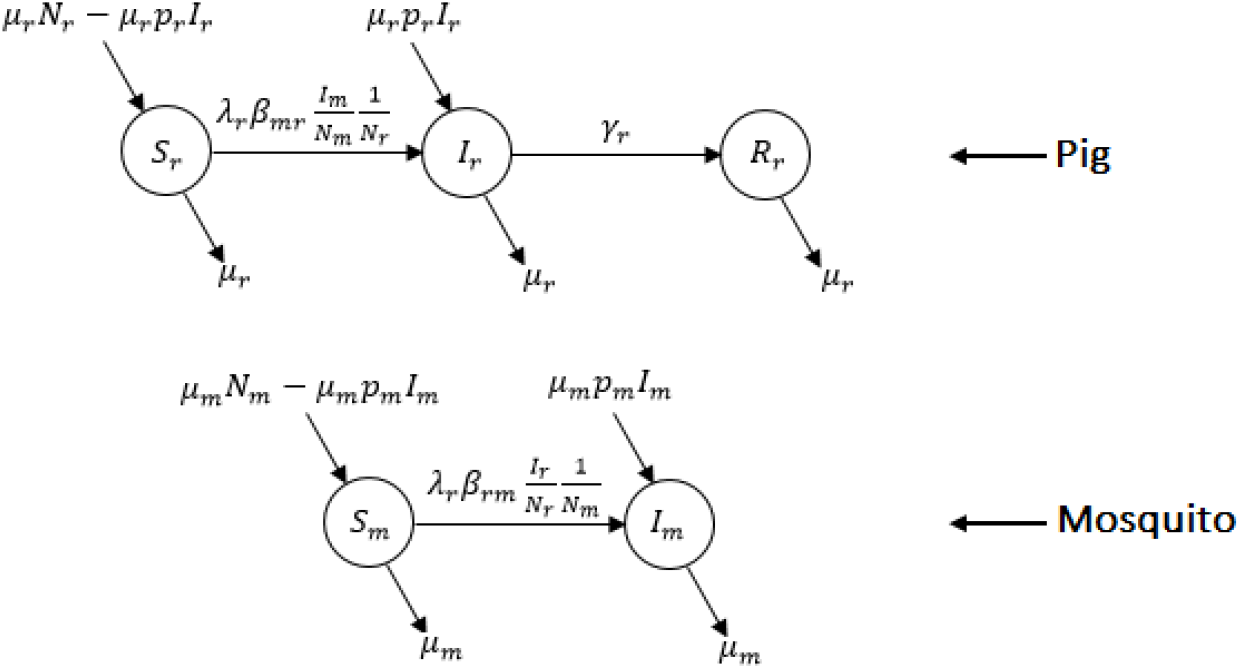
Flow diagram for pig and mosquito population.

**Figure 2:**
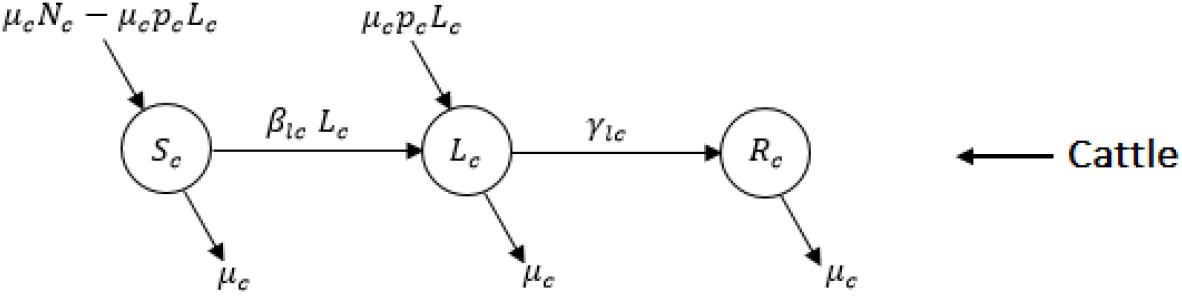
Flow diagram for cattle population.

**Figure 3:**
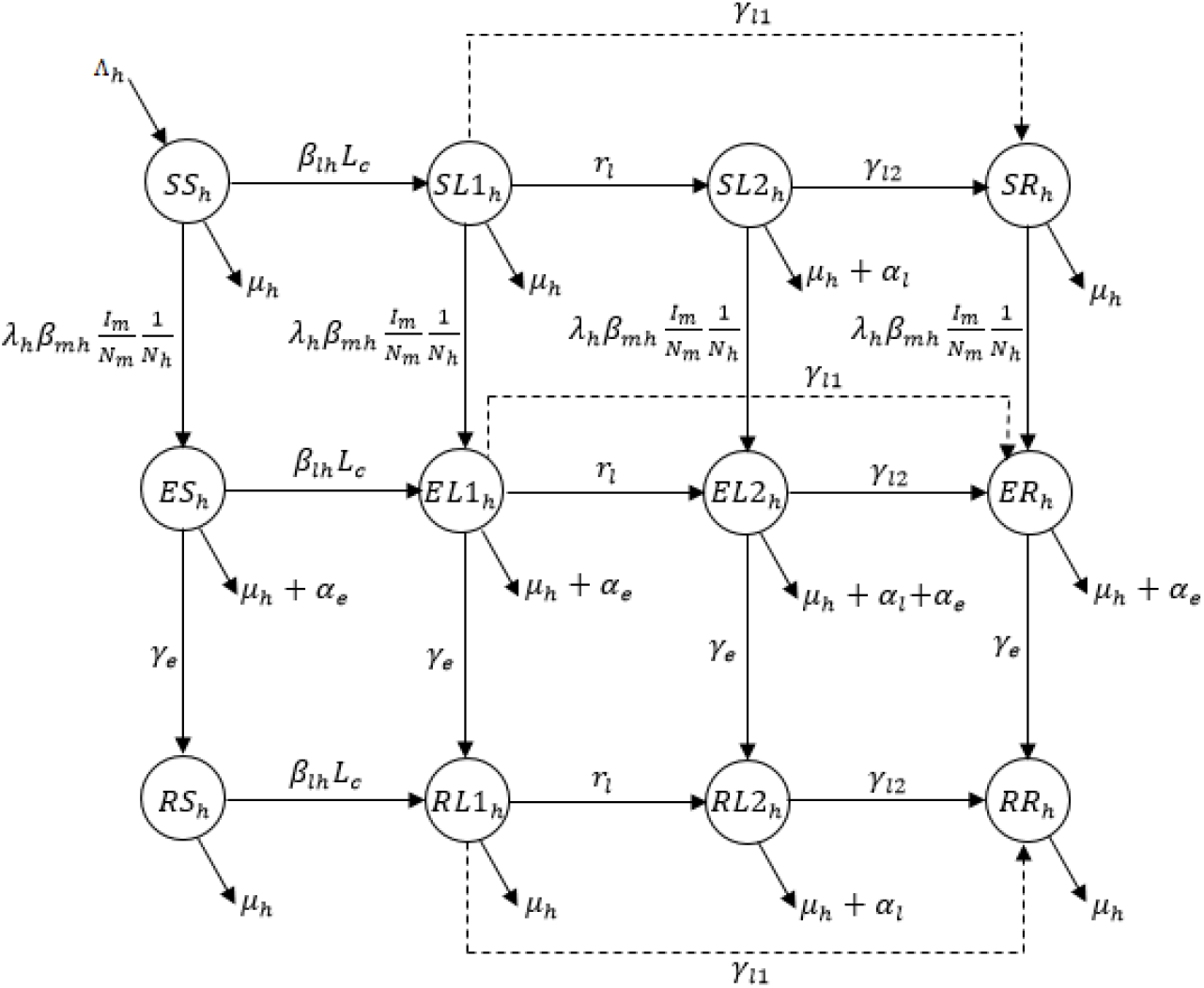
Flow diagram for human population.

Leptospirosis spreads through the urine of infected cattle or pigs and is transmitted through the contact of contaminated (by the urine of infected animal) surfaces or water [7]. Here, we ignore leptospirosis infections through pigs, because people usually keep pigs in a separate place from houses. Another important reason is its population size (*N*_*r*_), which is very small compared to the cattle population size (*N*_*c*_). Therefore, in our model cattle are the only source of *Leptospira*, which causes the disease. Hence, cattle and humans are getting infections just due to the presence of cattle (Figure 2 and Figure 3). Here, we consider that *Leptospira* can be transmitted in cattle through vertical transmission also.

Susceptible humans get leptospirosis infections and move to *SL*1_*h*_ from *SS*_*h*_. Most leptospirosis infected humans recover from the first phase, while the remaining move to the second phase of the infection (*SL*2_*h*_). Finally, patients from the second phase move to *SR*_*h*_ upon their recovery. The *SR*_*h*_ compartment represents people who already recovered from leptospirosis infection; however, they are still susceptible to JEV. A portion of susceptible humans get JE infections by infected mosquito bites before leptospirosis infection and move to *ES*_*h*_. Some of these infected humans get leptospirosis infection before their recovery from JE and move to *EL*1_*H*_. The remaining population of *ES*_*h*_ move directly to the JE recovered class (*RS*_*h*_). Similar to *SL*1_*h*_, most people from *EL*1_*h*_ move to *EL*2_*h*_ while others move to *ER*_*h*_. Recovered people from JEV infections (*RS*_*h*_) get leptospirosis infections and move to *RL*1_*h*_. Then, similar to *SL*1_*h*_ and *El*1_*h*_, most of them move to *RR*_*h*_ after recovery from leptospirosis, while others enter the second phase of infections (*RL*2_*h*_). Finally, people recover from *RL*2_*h*_ and move to *RR*_*h*_, which contains people who recovered from infections of both diseases.

In our model development, we consider constant populations for cattle, pigs, and mosquitoes. For disease transmission, we choose standard incidence for JEV, because infected mosquitoes (*I*_*m*_) are free to bite any individual among our host populations. On the other hand, we use mass-action incidence for *Leptospira* transmission, because the new infections depend on the availability of infected cattle (*L*_*c*_). Also, we assume our cattle, pig and mosquito populations to be constant.

## 3 Parameter estimation

The World Bank recorded the life expectancy of people at birth in India in 2017 as 69.165 years [28]. Taking the reciprocal of this value we get the natural death rate for humans as *µ*_*h*_ = 3.96*×*10^*−*5^*/*day. As per the 2014 census by the local health department, Malkangiri district had a population of 641,385 in 2014 with 109,483 households [29]. However, according to the World Bank data rural population in India in 2014 and 2018 was 876,035,725 and 892,321,651 respectively [30]. We use the ratio of these two populations to estimate the population of Malkangiri district in 2018, which gives us the population as 653,309 (*N*_*h*_) with 111,518 households. In our literature review, we did not find any value for the human recruitment rate. Hence, we use one of our equations (9th in the system) in a disease-free state which gives the equation Λ_*h*_ = *µ*_*h*_*N*_*h*_. Then we use this equation, and our already estimated values of *µ*_*h*_ and *N*_*h*_, to get Λ_*h*_ = 25.87 human*/*day.

We did not find any documented data for the size of cattle and pig populations (*N*_*c*_ and *N*_*r*_ espectively) in Malkangiri district. However, we found one documented data source which mentioned that the numbers of humans, cattle and pigs in Korkunda block of Malkangiri district in 2014 were 143,867, 80,583 (75,772+4,811), and 10,007 respectively in 29,667 households [22]. We use these numbers to estimate cattle and pigs per household, and then use our estimated number of households (111,518) to calculate numbers of total cattle and total pigs in the district in 2018, which gives 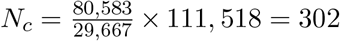, 911 and 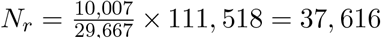. Now we need to know the feeding pattern of JEV vectors (value of *k*_*c*_ and *k*_*r*_) to estimate *λ*_*r*_ and *λ*_*h*_. We found a study that estimated the feeding preference for JE vectors in the southern part of India as 46.4% on cattle, 4.8% on pigs, and 1.5% on humans [31]. Using these values we have 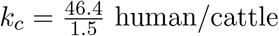 and 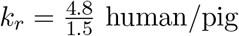.

To estimate *λ*_*r*_ and *λ*_*h*_, we also need to have the size of JEV carrier mosquito populations (*N*_*m*_), along with their biting rate (*b*_*m*_). A field study in rural villages of Western Yunnan Province of China was done in 2013 to estimate the abundance of mosquitoes in an Asian rural setting [32]. Researchers collected mosquitoes from two households in each of four studied villages in each month for a 12 month-study. The total collection of mosquitoes was 85,307. It is equivalent to 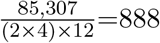 mosquitoes/household. So, for our study we have a total of (111,518×888)=99,027,984 mosquitoes. Another study carried out in some villages of Malkangiri district showed that 69.1% of available mosquitoes are vectors for JEV [22]. Hence, the total number of JEV vectors we have is *N*_*m*_=99,027,984*×*69.1%=68,428,337. We found documented data for the biting rate of JEV vectors which give *b*_*m*_=0.25 bite/mosquito-day (mean of 0.2 and 0.3) [33]. Anyway, not all mosquito bites belong to these three hosts. Also, blood-meal analysis showed that some mosquitoes bite more than one host [31]. Hence, we estimated that about 70% of mosquito bites are distributed among cattle, pig and human population. Thus, 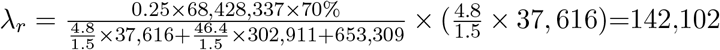 bite (total)/day and 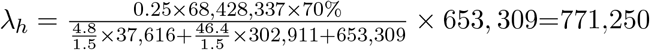 bite (total)/day.

In 2000, the Indian Council of Medical Research carried out a study which found the transmission and infection rate for JEV vectors [34]. The study showed that vectors have 82% infection probability infection probability from pigs; however, the transmission probabilities of infections from mosquitoes to humans and mosquitoes to pigs are 31.6% and 63.5% respectively. Hence, we have *β*_*rm*_ = 0.82 mosquito*/*bite, *β*_mr_ = 0.635 pig*/*bite, and *β*_mh_ = 0.316 human*/*bite. A separate review study the expansion of JEV carried out in 2009 which mentioned that pigs maintain enough viremia to infect mosquitoes for up to 4 days [36]. Taking the reciprocal of this value we get 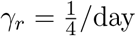. For JE vectors’ mortality rate, we found a study that estimated their life expectancy as 59.8 days [37], and this gives us 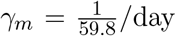. Humans’ mean recovery period during the 2012 JE outbreak in Odisha was 7 days [35], which gives us 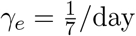. For the estimation of *µ*_*r*_, we found that domestic pigs have an average lifespan of 6-10 years, but, that can be shorter due to certain problems [38]. Thus, instead of the median (8 years) of the range, we choose 7 years as the life expectancy for pig population which eventually estimated 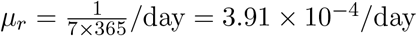.

A review study from 2014 mentioned the mean durations for the acute phase (1st phase, in our model) and immune phase (2nd phase, in our model) as 10 days and 14 days respectively [39]. Taking reciprocals of these values we get 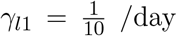 and 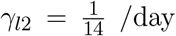. Another study in coastal south India estimated that 51 out of 202 (25.24%) leptospirosis patients move to the 2^nd^ phase [40]. Hence, we use this result and the ratio 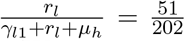 to estimate the transfer rate from phase 1 to phase 2, and found *r*_*l*_ = 0.033**/day**. We found a couple of documented data for cattle recovery period of leptospirosis which are inconsistent in values [41, 42, 43]. Hence, based on those available values we choose that cattle can shed *leptospira* in their urine for 60 weeks, which gives 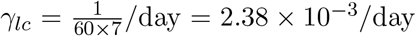, a different disease in cattle in Odisha state was studied which had the highest cattle age group of 6.5-7.5 years; however, they did not mention the highest or average age [44]. So, we take 8 years as the lifespan for cattle in Odisha, which gives us 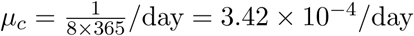.

We did not find enough evidence for vertical transmission of leptospirosis in cattle (*p*_*c*_) and of JEV in pigs (*p*_*r*_), and therefore we assume *p*_*c*_ = 0 and *p*_*r*_ = 0. However, we found 4% effective vertical transmission for JEV vectors, which gave *p*_*m*_ = 0.04 [45]. In 2016, in Malkangiri district 37 died out of 175 JE patients [29]. Using this ratio and the relation 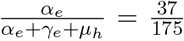, we estimated *α*_*e*_ = 0.0383*/*day. The case fatality rate for leptospirosis infection is 7.69% [50]. Also, we already know that about 25.24% leptospirosis cases move to the 2nd phase [40]. Hence, the case fatality rate among patients of 2nd phase is 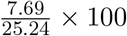, which implies *α*_*l*_ = 3.13 *×* 10^*−*2^/day.

During our literature review, we did not find any value for infection rates of leptospirosis in cattle and in humans (*β*_*lc*_ and *β*_*lh*_ respectively). Hence, we follow the method used in [20, 19] to estimate *β*_*lc*_ and *β*_*lh*_ which gives 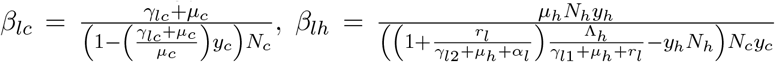 where and *y*_*h*_ are prevalence of leptospirosis in cattle and in humans respectively. We did not find any documented data for *y*_*c*_ in Malkangiri. However, we found one study from 2013 which estimated leptospirosis prevalence as 20.7% in cattle in a neighbour district of Malkangiri [27]. We choose 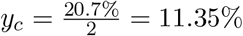 since the sample collections of this study were not random, rather were mostly from the villages with a history of abortions and other disorders. Also, we did not find documented data for leptospirosis prevalence in humans in Malkangiri. However, we found human and cattle prevalence of leptospirosis in another prevalent location of leptospirosis, northeast Thailand. We estimated leptospirosis prevalence in cattle for this area to be 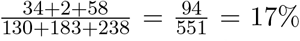 using the data from a study published in 2016 [47]. A different study estimated 12.5 annual cases of human leptospirosis per 100,000 people [48]. We used the ratio of estimated leptospirosis prevalence in cattle for our study area (11.35%) and of northeast Thailand (17%), and the annual cases of human leptospirosis in northeast Thailand (12.5 per 100,000) to calculate annual cases of human leptospirosis for our study area, and found 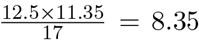 cases per 100,000 people, which is equivalent to 0.00835%. Also, we have a weighted recovery period as 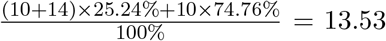 days, which gives us 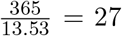 as the number of generations in a year for infected people. Finally, we got 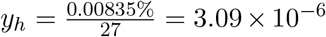. Using these prevalence values and our expressions for leptospirosis infection rates, we got *β*_*lc*_ = 9.66 *×* 10^*−*8^/day-cattle and *β*_*lh*_ = 9.11 *×* 10^*−*12^/day-cattle.

## 4 Analysis

We begin with the general model where we consider the presence of cattle which includes both JE and leptospirosis diseases. Later, we consider the scenario without cattle; this special scenario has JEV infection only. Finally, we estimate total disease burdens for these two scenarios and compare them to understand the impact of cattle on disease burden in JE prevalent areas. The goal of this study is actually to understand how the presence of domestic animals, here cattle, helps to reduce disease burden.

First, we analyse the cattle system, which is decoupled from the other two subsystems – the pig-mosquito system, and the human system. We set all the equations of this system equal to zero and solve them for state variables to find equilibria values. After performing some basic algebra we get the disease free equilibrium (*DFE*) as *S*_*c*_ = *N*_*c*_, *L*_*c*_ = 0, *R*_*c*_ = 0, and the endemic equilibrium (*EE*) as

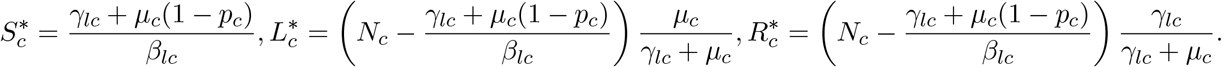

Then we use the next generation method [49] to obtain the basic reproduction number (*BRN*) which gives 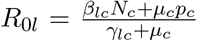. We find that the *EE* exists under the condition *R*_0*l*_ *>* 1. Here the *EE* is globally asymptotic stable if and only if *R*_0*l*_ *>* 1, otherwise the *DFE* is globally stable (see appendix 1 for details).

Before we begin our discussion on the pig-mosquito subsystem, here we consider a simplifying assumption that the two diseases do not significantly affect the magnitude of the human population, so that we can replace *N*_*h*_(*t*) by *N*_*h*_(0) in the denominator of *λ*_*r*_ and *λ*_*h*_, in order to simplify the analysis. This assumption is supported by both our literature review and the later numerical analysis. As per the study [29], JEV infection 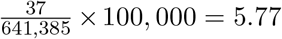 annual deaths per 100,000 people. Also, leptospirosis infection causes 12.5 *×* 7.69 = 0.96 annual death per 100,000 people [48, 50]. These two estimations give us a total of 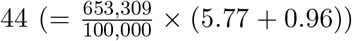 annual deaths due to both diseases, which is 0.0067% of the total human population. Under this assumption, mosquito’s biting rates to pigs, and humans (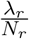 and 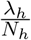 respectively) become constants, and the vector-reservoir (pig-mosquito) subsystem is decoupled from the human subsystem. The pig-mosquito subsystem is already decoupled from the cattle subsystem. Now, following the same approaches, like the cattle system, for the pig-mosquito system, we get the *DFE* as *S*_*r*_ = *N*_*r*_, *I*_*r*_ = 0, *R*_*r*_ = 0, *S*_*m*_ = *N*_*m*_, *I*_*m*_ = 0, and the *EE* as

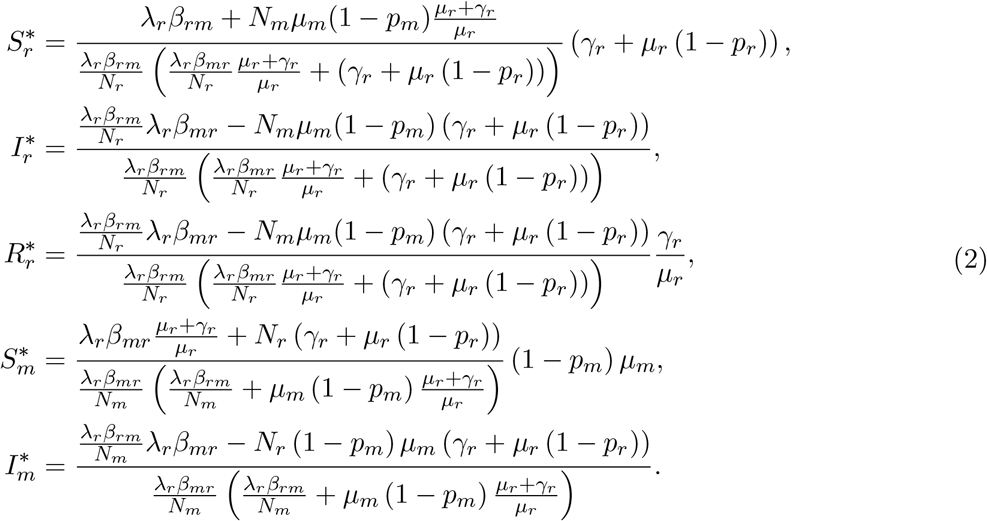

We get the *BRN* for the pig-mosquito system as

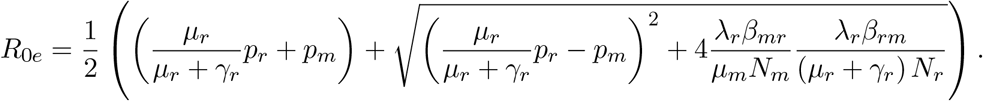

Here the *EE* exists and is locally asymptotically stable (LAS) if and only if *R*_0*e*_ *>* 1, otherwise the *DFE* is LAS. Next, we develop a strong Lyapunov function to show that the *DFE* is globally stable when *R*_0*e*_ *<* 1 (see appendix 2 for details). Then, we analyse the stability of the *EE* numerically which indicates that the *EE* here is globally stable if and only if *R*_0*e*_ *>* 1.

The cattle subsystem and the pig-mosquito subsystem are decoupled from the human subsystem respectively by the model and the assumption that the magnitude of the human population is insignificantly affected by the effect of JE and leptospirosis. As these two subsystems go to equilibrium, the limiting system for system 1 is given by the human subsystem with *L*_*c*_ and *I*_*m*_ replaced by their equilibrium values (please see system (3)). Here *N*_*m*_ is constant which makes the term 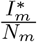 to be constant and 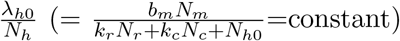 is also constant. Thus, the resulting system (system (3)) is linear and has only one equilibrium (see equation (9) in Appendix 3) where the equilibrium values are functions of 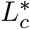 and 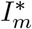 Hence, the behavior of the limiting system follows the behavior of our cattle and pig-mosquito subsystems. Now, a theorem by Horst R. Thieme [51] leads us to the fact that the behavior of the entire system is asymptotic to the behavior of the limiting system, which is linear. Eventually, the entire system is governed by the behavior of the cattle and the pig–mosquito subsystems.

Next, we extend our analysis to the entire system, which includes the cattle, the pig–mosquito, and the human subsystems. We obtain the invasion reproduction number (*IRN*) for both diseases following the approach by Mitchell & Kribs [52]. Each of these *IRN* s is found to be the same as the corresponding *BRN* s. Thus, both of the *IRN* s are independent of the presence of the other disease. This result is easily comprehensible since neither of these diseases’ transmissions is affected by the other. Our preceding analyses found two different values for both 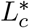 and 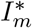; hence, their all possible combinations produces four different equilibria for the entire system. These four equilibria are – *DFE*, endemic in leptospirosis and free of JE, endemic in JE and free of leptospirosis, and endemic in both diseases respectively. So, for the entire system, we have four different scenarios – (i) *R*_0*l*_ *<* 1 and *R*_0*e*_ *<* 1 (ii) *R*_0*l*_ *>* 1 and *R*_0*e*_ *<* 1, (iii) *R*_0*l*_ *<* 1 and *R*_0*e*_ *>* 1, and (iv) *R*_0*l*_ *>* 1 and *R*_0*e*_ *>* 1.

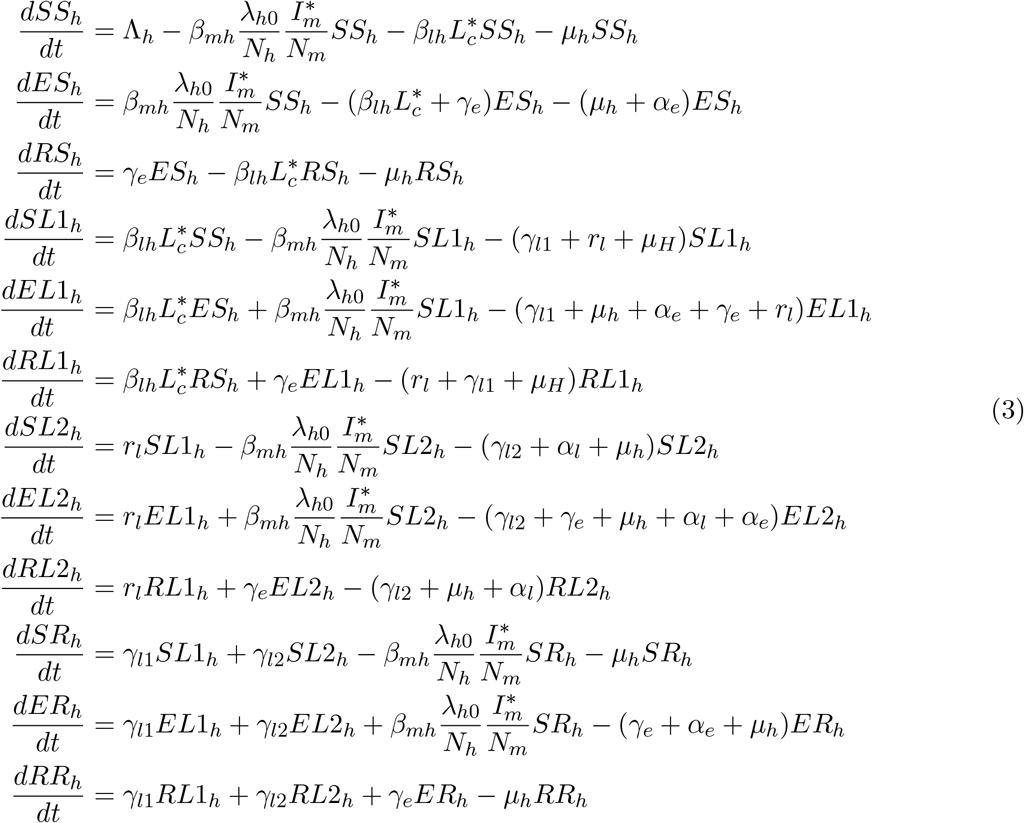

where 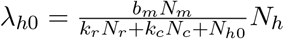 (here *N*_*h0*_ = *N*_*h*_(0)).

Now we consider the scenario without cattle which eliminates the possibility of leptospirosis infections in humans. Here the *BRN* for JE (*ℛ*_0*e*_) is different compared to the *BRN* (*R*_0*e*_) from from the general model since the *λ*_*r*_ in *R*_0*e*_ is a function of cattle population size. The absence of cattle reduces the human system to a three-dimensional system, which gives the *EE* for the human system as

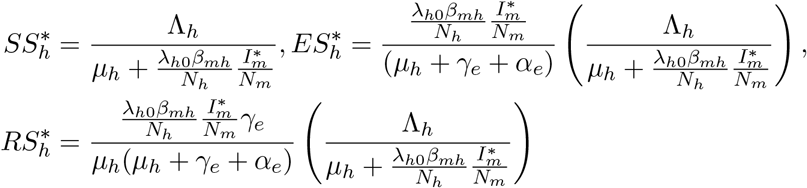

where 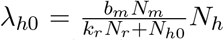 (here *N*_*h*0_ = *N*_*h*_(0)). This special case has *DFE*, and the only endemic equilibrium (*EE*) when *ℛ*_0*e*_ *>* 1. This *EE* is completely governed by 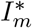 (see equation (2) for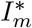).

To better understand the impact of cattle, some numerical analyses are done based on our parameter estimations. Using our estimated parameter values, we find *R*_0*l*_ = 10.35 and *R*_0*e*_ = 1.008 respectively in the presence of cattle. However, our estimations in the absence of cattle found *ℛ*_0*e*_ = 12.97. Based on our estimated parameter values, in the presence of cattle we estimate 72 and 228 annual cases for leptospirosis and JE respectively, whereas the annual deaths are 6 and 48 respectively. In the absence of cattle, we estimate 9,407 and 1,988 number of annual cases and annual deaths due to JEV infections. Next, we perform numerical analysis to estimate total cases and the BRN of JE and leptospirosis as the average number of cattle per household varies and plot the related graphs (Figure 4). The Figure 4(a) shows that the JEV infection decreases as the number of cattle increases; however, the human leptospirosis incidence increases with the cattle; the Figure 4(b) illustrates that BRN for JE exponentially decreases, and for leptospirosis linearly increases as the number of average cattle increases. So, the presence of cattle has positive and negative impacts on human health.

**Figure 4:**
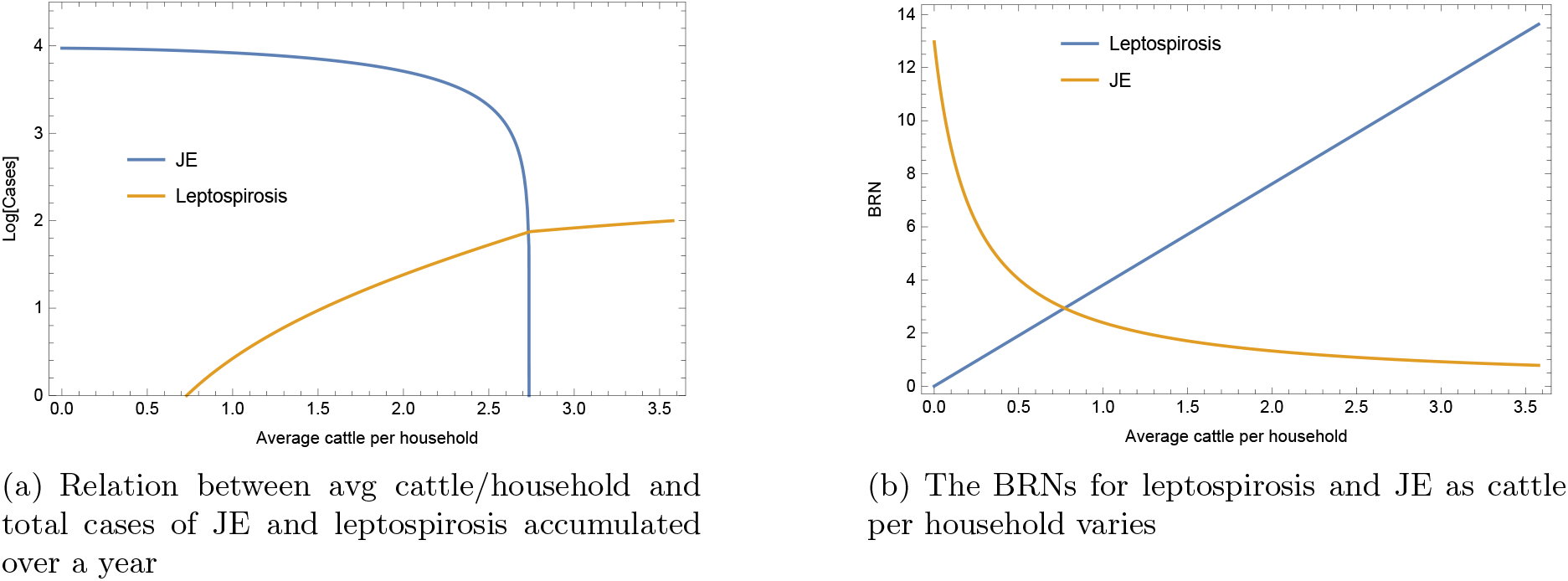
The effect of cattle on infections and BRNs.

To quantify the impact of cattle on human health, we calculate the total disease burden for our two settings – with cattle and without cattle. In this calculation we follow an approach similar to the approach of the study [53] where they followed WHO guidelines [54]. To calculate the total disease burden they estimated total *DALY* (Disability-Adjusted Life Year) where one *DALY* means the loss of one year of healthy life. The *DALY* is composed of *Y LD* (Years Lost due to Disability) resulting from infections and *Y LL* (Years of Life Lost) caused by disease-induced premature deaths which are calculated using formulas *Y LD* = *I × DW × L*_1_ and *Y LL* = *D × L*_2_, where *I* and *D* represent the total number of infections (cases) and total number of deaths respectively. Here *L*_1_ = average duration (in years) of illness which is the reciprocal of the recovery rate for survival cases and of the death rate for non-survival cases, and *L*_2_ = standard life expectancy at age of death (in years) = average life expectancy of the community – average age at premature death and the term DW is the disability weight for diseases which ranges from 0 (perfect health) to 1(death). It can be thought of as the proportional reduction in perfect health due to any adverse health condition caused by infections. So, the total burden of disease can be represented as the sum of *Y LD* and *Y LL*, which gives

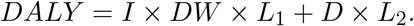

In this work, we study two different diseases, JE and leptospirosis. Hence, we will have different values of *I, D, L*_1_, *L*_2_ and of *DW* for JE, and leptospirosis. Here the quantities *L*_1_s are the reciprocals of the *γ*_*l*1_, *γ*_*l*2_ for the first and second phases of leptospirosis respectively, and the reciprocal of the *γ*_*e*_ for JE. To estimate *L*_2_s, we need to know the average ages of infections which are 40.48 years [40] and 5 years [35] for leptospirosis and JEV infections respectively. The average age of infections and the average age of deaths are the same for leptospirosis; however, they are different for JE (5 years and 3 years respectively [35]). The values of *DW* differs between leptospirosis and JE; it also differs between phases of leptospirosis. Furthermore, the *DW* has different values for survival and non-survival. All these different values of *DW* are taken from the study [55] based on the severity (mild or severe) of illness and on the onward complications upon recovery. Therefore, using the above formula for *DALY* we calculate total *DALY* s separately for JE and leptospirosis infections and add them to have total *DALY* cause from both diseases combined.

Based on our parameter estimations, the above formula and values together calculate 4,396 *DALY* s and 157,642 *DALY* s for the setting with cattle and without cattle respectively. These results clearly show that the presence of cattle has a huge positive impact on the disease burdens in JE-prevalent areas. Next, we calculate total *DALY* s varying the average number of cattle per household to understand how the variation in the average number of cattle per household changes the annual disease burden. The results of these calculations are portrayed in Figure 5, which evidently illustrates that the presence of cattle is unconditionally helpful in terms of the annual disease burden. However, there is an optimum value for the average number of cattle per household to ensure the minimum disease burden which is 2.75. Maintaining cattle beyond this threshold value the annual disease burden increases slowly which is generated from the infections of leptospirosis. On the other hand, keeping fewer cattle per household than the optimum number causes a sharp rise in the annual disease burden due to a significant increase in JE infections.

**Figure 5:**
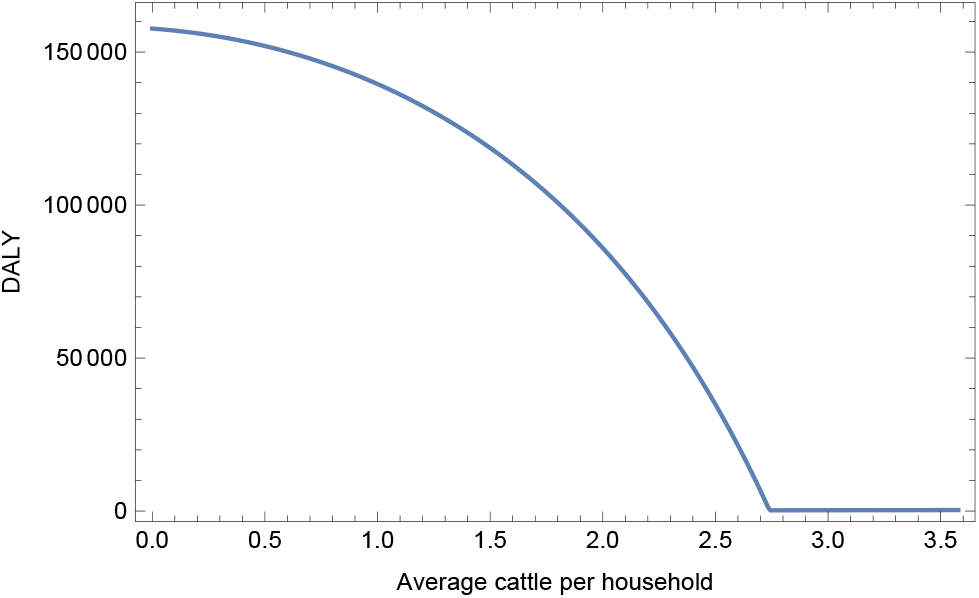
Relation between avg cattle/household and annual disease burden (in *DALY* s)

## 5 Discussion

To the best of our knowledge, this is the first work to understand the joint dynamics of JE and leptospirosis. Our qualitative analysis exhibits the classic threshold behavior of the system. The quantitative analysis shows that the presence of cattle reduces total disease burden even though it introduces leptospirosis. It happens because the number of JE cases is reduced by the cattle presence at a higher rate than the rate at which humans get leptospirosis infections due to the presence of cattle. Also, the case fatality rate (CFR) of JE infections is higher than the CFR of leptospirosis infections. The disability weight is higher for JE infections than for the leptospirosis infections, and the average age of JEV infections is very low compared to the average age of leptospirosis infections. Even though the presence of cattle is always helpful, there is an optimum average number of cattle per household which minimizes the annual disease burden.

Our analysis found that the presence of cattle helps to reduce the annual disease burden in JE-prevalent areas because of the vector’s (mosquitoes’) feeding preference. Mosquitoes prefer to bite cattle at a much higher proportion compared to the proportion to bite humans or pigs. Thus, cattle reduce mosquito-human interactions and eventually reduce human cases of JEV infections. This result is consistent with the result of an ecological study by Miller and Huppert [17] where they identified that vectors’ feeding preference decides whether or not host richness is helpful to dilute disease prevalence. Also, our results found that the JE prevalence in humans decreases as the cattle population increases. This finding is consistent with the finding of another ecological study on vector-borne diseases by Johnson and Thieltges [16] where they found that the strength of the dilution effects depends on the relative abundance of the dilution host to the focal host.

This study didn’t consider seasonality in its model development. Considering seasonality will provide a more accurate picture of short-term dynamics. We didn’t include seasonality because the concern of our study is primarily with the overall disease burden, not with the detailed disease dynamics. In our analysis, we assumed constant biting rates to pigs and humans which should not have any significant impact on our results since the mosquito population is not limited by the available host populations, rather by available breeding sites. We also assumed a homogeneous distribution among households and those cattle close enough to households to affect the infection risks for JEV and leptospirosis. Large cattle farms farther away from homes than pigs are, are not likely to affect those risks (except to employees). However, studies have shown that heterogeneity does affect disease risk [56, 57]. In our case, heterogeneity is the heterogeneous distribution of household cattle per household.

## Data Availability

All used data are included with reference in the manuscript.

## Appendix 1 Stability of the endemic equilibrium of the cattle system

### Proposition 1

*The endemic equilibrium for the cattle subsystem is locally stable if and only if R*_0*l*_ *>* 1.

Proof: We find the Jacobian matrix of the cattle system at the *EE* which gives the characteristic equation

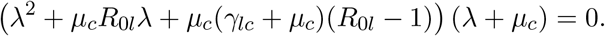

Here, *λ* = *−µ*_*c*_ is negative. Next, we use Routh-Hurwitz Criteria to check if the real part of remaining eigenvalues are negative. As per Routh-Hurwitz Criteria, the coefficients of *λ* and the constant term need to be positive to ensure the negativity of real parts of eigenvalues. Here it is evident that the coefficient of *λ* is always positive and the constant term is positive when *R*_0*l*_ *>* 1. Hence, the *EE* for the cattle subsystem is locally stable if and only if *R*_0*l*_ *>* 1.

### Proposition 2

*The endemic equilibrium for the cattle subsystem is globally stable when R*_0*l*_ *>* 1, otherwise the *DFE* is globally stable.

Proof: The first two equations of the cattle subsystem are

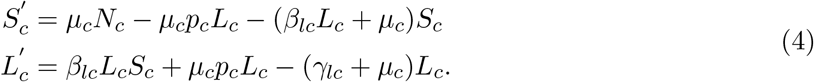

It is evident from the system (4) that 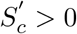 if *S*_*c*_ = 0 and 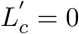 if *L*_*c*_ = 0, which means the system (4) is well posed. Now, the system (4) gives 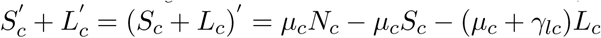 From this relation it is evident that (*S*_*c*_ + *L*_*c*_)′ ≤ 0 if either *S*_*c*_ ≥ *N*_*c*_, or *L*_*c*_ ≥ *N*_*c*_ which shows all solutions are bounded.

Now we define 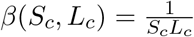 to use Dulac’s Criterion to check if the *EE* approaches a limit cycle. Here we assume 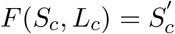 and 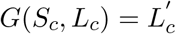 which give

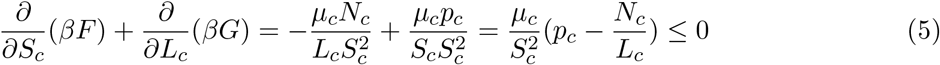

So, no limit cycle exists.

Hence, by Poincaré-Bendixson Theorem all solutions approach the *EE* when *R*_0*l*_ *>* 1 which means the *EE* is globally stable if and only if *R*_0*l*_ *>* 1, otherwise the *DFE* is globally stable.

## Appendix 2 Stability of the endemic equilibrium of the pig-mosquito system

### Proposition 3

*The endemic equilibrium for the pig–mosquito subsystem is locally stable if and only if R*_0*e*_ *>* 1.

Proof: We can reduce our system to a system of three equations since pig and mosquito populations are constant. We Replace *S*_*r*_ = *N*_*r*_ *−* (*I*_*r*_ + *R*_*r*_) and *S*_*m*_ = *N*_*m*_ *− I*_*m*_ in the 3rd and 5th equation of the system (1) and get the reduced pig-mosquito system as

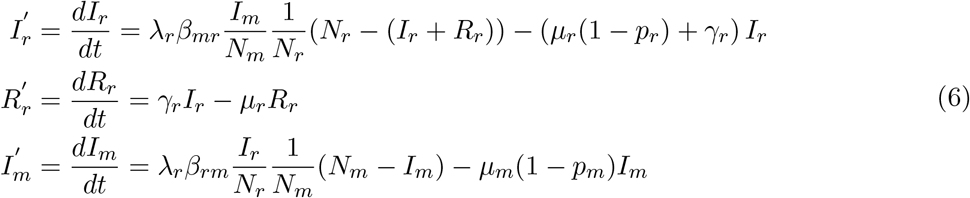

Next, we find the Jacobian matrix of the reduced system (6) at the *EE* which gives the characteristic equation

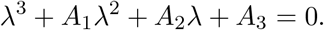

Here, *A*_1_ = *−*(*a*_1_ + *a*_2_ + *a*_3_), *A*_2_ = *a*_1_*b*_2_ *−a*_2_*b*_1_ + *a*_1_*c*_3_ *−a*_3_*c*_1_ + *b*_2_*c*_3_, and *A*_3_ = *a*_3_*b*_2_*c*_1_ + *a*_2_*b*_1_*c*_3_ *−a*_1_*b*_2_*c*_3_ where 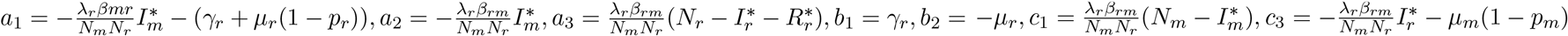.

Next, as per Routh-Hurwitz Criteria, we verify *A*_1_, *A*_2_ *>* 0 and *A*_1_*A*_2_ *> A*_3_ to check if the real parts of all eigenvalues are negative. Here, *A*_1_ is clearly positive. Also, our analysis found *A*_2_ *>* 0 and *A*_1_*A*_2_ *> A*_3_.

Hence, the endemic equilibrium for the pig–mosquito subsystem is locally stable if and only if *R*_0*e*_ *>* 1.

### Proposition 4

*The DFE for the pig–mosquito subsystem is globally stable when R*_0*e*_ *<* 1.

Proof: We define our Lyapunov function as

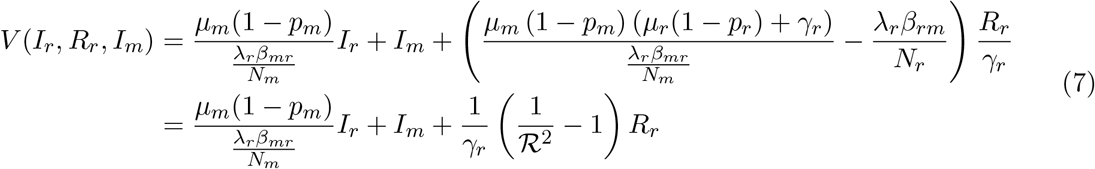

which gives

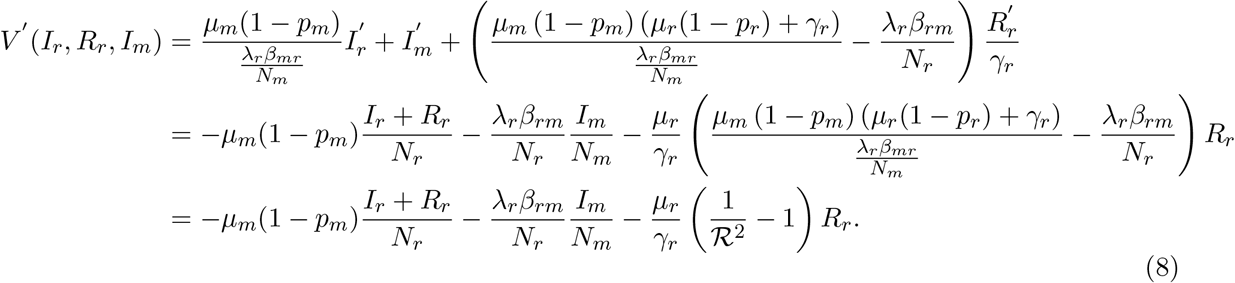

Here we use a alternative threshold quantity *R* to understand if the function *V* satisfies all the conditions of a strong Lyapunov function which is defined as

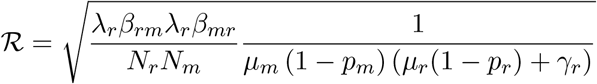

where *R >* 1 if and only if *R*_0*e*_ *>* 1.

Now, the alternative threshold value *R*, equation (7) and equation (8) easily verify that

*V* (0, 0, 0) = 0 and *V* (*I*_*r*_, *R*_*r*_, *I*_*m*_) *>* 0 for (*I*_*r*_, *R*_*r*_, *I*_*m*_) ≠ (0, 0, 0) when *R <* 1.

Also, *V* (0, 0, 0) = 0 and *V* (*I*_*r*_, *R*_*r*_, *I*_*m*_) *<* 0 for (*I*_*r*_, *R*_*r*_, *I*_*m*_) ≠ (0, 0, 0) when *R <* 1.

Hence, the *DFE* for the pig-mosquito system is globally stable when *R*_0*e*_ *<* 1 (since *R <* 1⇔ R_0*e*_ *<* 1).

## Appendix 3. Equilibrium of the limiting system

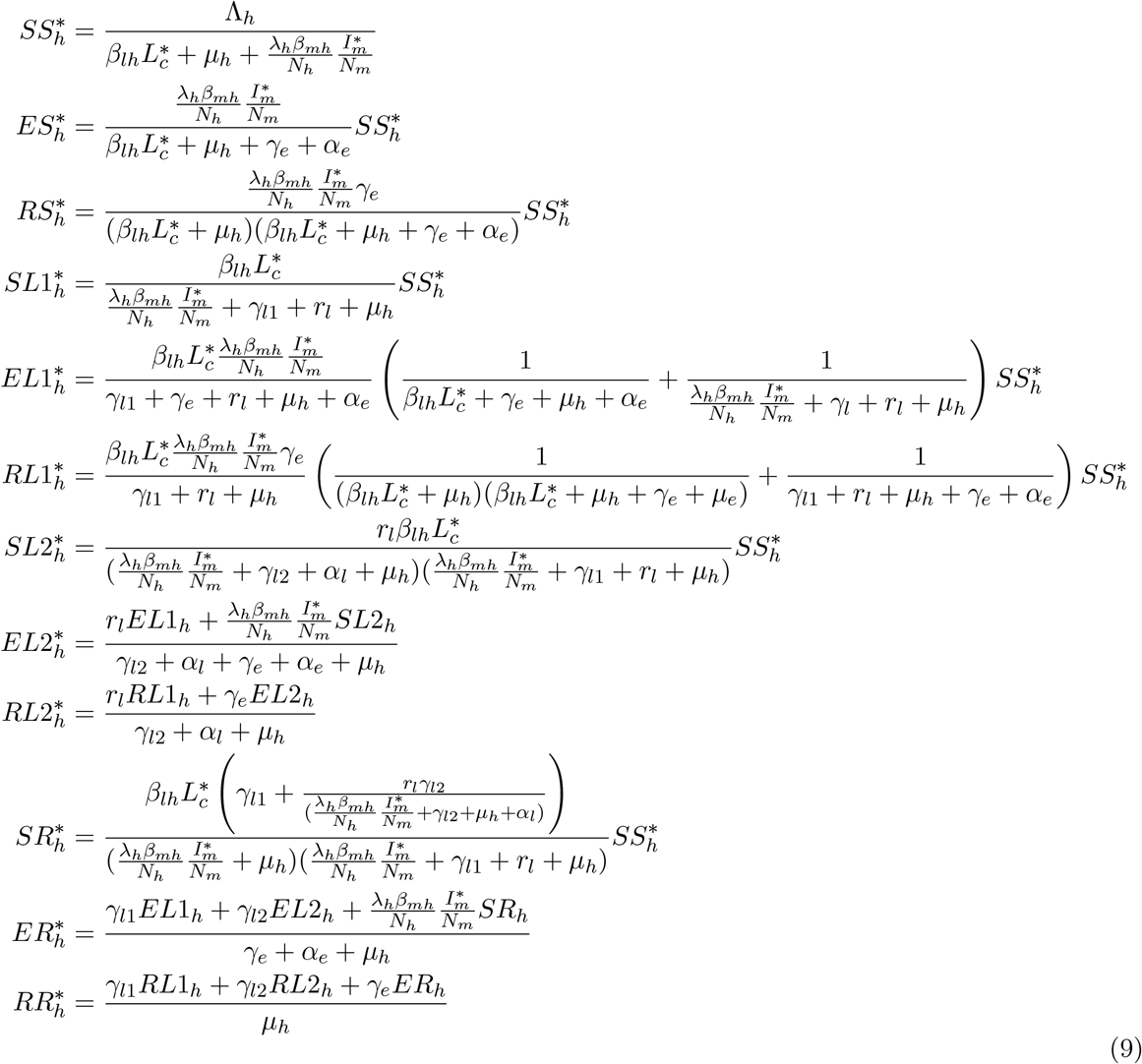

